# Racially Focused LGBQ Pride Events: The Promotion of Psychological Wellbeing Among Cisgender LGBQ People of Color

**DOI:** 10.1101/2022.07.13.22277611

**Authors:** Michael G. Curtis

**Author notes:** Corresponding author: Michael G. Curtis, University of Georgia, 1095 College Station Road, Athens, Georgia 30602-4527.

## Abstract

Lesbian, gay, bisexual, and queer/questioning people of color (LGBQ POC) are disproportionately more likely to experience issues related to their mental health when compared to their White counterparts. In spite of this persistent mental health disparity, few studies have been dedicated to identifying mental health-related promotive factors among LGBQ POC adults. The current study examined the extent to which attending LGBQ POC Pride events (e. g., Black Pride Festival) was associated with cisgender LGBQ POC’s psychological wellbeing. I hypothesized that attendance would be positively associated with psychological well-being due to the unique sociocultural resources offered at these events. I further hypothesized that participants’ racial/ethnic identity or gender would modulate these effects. Hypotheses were tested using multiple linear regression with data from the 2013 Social Justice Sexuality Project (n = 2,486). Attending LGBQ POC Pride events was positively associated with cisgender LGBQ POC’s psychological wellbeing. Post hoc multigroup analysis indicated that participants’ racial/ethnic identity or gender did not moderate these effects. Findings suggest that attending racially focused LGBQ Pride events may offer cisgender LGBQ POC’s unique psychological resources that are not provided by general LGBQ Pride events.

**Public Health Significance:** LGBQ people of color (POC) are a multiply marginalized population vulnerable to increased levels of psychological distress that is unquietly different from their White LGBQ counterparts. Findings suggest that increasing the prevalence of racially focused Pride events may contribute to the psychological wellbeing of LGBQ POC.

## Introduction

Empirical research has consistently demonstrated that lesbian, gay, bisexual, and queer/questioning (LGBQ) adults are at a higher risk of developing issues related to their mental health, when compared to their heterosexual counterparts. ^1-3^ One such study demonstrated, with the use of epidemiological data from the United States and United Kingdom, that rates of depressive, anxiety, and substance use disorders are 1.5 to 3.0 times higher in among LGBQ adults. ^4^ Studies dedicated to investigating mental health related promotive factors among LGBQ adults are scarce in spite of evidence of persistent mental health disparities. ^5,6^

Disparities in mental health among LGBQ people, compared to their heterosexual counterparts, are often explained by the minority stress model. ^7^ The minority stress model posits that sexual minorities are at increased risk for negative mental health outcomes due to their exposure to unique stressors related to their sexual minority identities, such as discrimination and victimization. ^8^ Consistent with this model, a large body of work suggests that experiences of social isolation, peer victimization, and family rejection are associated with significant reductions in psychological wellbeing in samples of LGBQ adults. ^9,10^ Among LGBQ people of color (POC), these disparities are exacerbated due to their distinctive experiences of both racism and heterosexism. ^11^ Nascent research suggests that the synergistic effects of these reinforcing forms of discrimination evidence adverse mental health outcomes above and beyond that of their White LGBQ peers. ^12-15^ Prior research indicates that LGBQ POC experience experiences greater stress, less support, and fewer resources than White LGBQ people. ^16,17^ Additionally, LGBQ POC are known to experience increased family rejection and a greater tendency to conceal their sexual orientation than White LGBQ adults. ^18,19^

In response, LGBQ mental health scholars have aimed to identify factors that are stress-ameliorating among LGBQ POC. For instance, social support has emerged as a consistent and robust promotive factor against psychological distress among LGBQ POC adults. ^20,21^ Prior research indicates that social support is effective in promoting psychological wellbeing among LGBQ POC because as it is associated with increases in sexuality self-acceptance, comradery among LGBQ POC of similar racial/ethnic backgrounds, and culturally responsive coping resources. ^22,23^ Social ecological research frameworks posit that social support factors at multiple social levels (including peers, family, organizations such as schools, communities, and society) may provide needed support and contribute to improved health behaviors and well-being. ^24,25^ Indeed, prior research indicates that attending community-level events, such as LGBQ Pride Events, may diminish the prevalence of these mental health disparities among LGBQ adults by (a) fostering a sense of communal support and (b) decreasing individuals’ experiences of social isolation. ^26-28^

The genesis of many LGBQ Pride events can be traced back to public protests and organized demonstrations dedicated to commemorating the Stonewall Riots of 1969. ^29^ Since then, the overall tone of Pride events has evolved from being focused on political advocacy to general celebrations of LGBQ communal progress and visibility. ^30^ As a result of this shift in tone, critical activists and scholars have posited that Pride events’ political, and especially the radical, aspects have been significantly diminished. ^30,31^ Although many activist groups continue to maintain the original spirit of Pride events by engaging in community building and advocacy efforts within the context of tourism-centered events. ^32,33^

The transformation of Pride events into celebratory spaces has expedited their commodification, often referred to as rainbow capitalism or pink tourism. ^34,35^ Puar ^36^ notes how the commercialization of Pride events has resulted in organizers privileging the wants, needs, and desires of white middle/upper-class gay men and lesbian tourists due to their increased ability to purchase goods and services. Consequently, prior evidence indicates that LGBQ community members who are representative of racially and ethnically diverse groups frequently report feeling unwelcome or uncomfortable at most LGBQ Pride events. ^37,38^

The feelings of unwelcomeness frequently experienced by LGBQ POC ignited a trend of organizations creating racially focused Pride events, such as Atlanta Black Pride or Celebrate Orgullo, Miami’s first Hispanic LGBT Pride festival. ^31,37,38^ In particular, many LGBQ POC report feeling as though their unique lived experiences were consistently overshadowed by larger cultural discourses within their racial-ethnic communities and the larger LGBT community, respectively. ^39,40^ Consequently, prior evidence indicates that attending racially focused LGBQ Pride events may cultivate areas of resilience among LGBQ POC by creating a distinctive, celebratory space that centers their unique experiences. ^39,40^ Despite converging evidence indicating the distinctive positive effect of attending racially focused LGBQ Pride events on LGBQ POC’s psychological wellbeing, no study, to date, has empirically examined this association.

## The Current Study

Framed by an intersectional perspective and informed by minority resiliency models, the current study aimed to examine the extent to which attending racially focused LGBQ Pride events was associated with cisgender LGBQ POC’s psychological wellbeing. First, I hypothesized that because of the unique sociocultural resources offered by attending racially focused LGBQ Pride events, attendance would be positively associated with psychological wellbeing above and beyond general feelings of LGBQ connectedness. I hypothesized that these effects would be modulated by an individual’s racial identity or gender. I expected that the association between attending racially focused LGBQ Pride events and psychological wellbeing would be more pronounced among Black Americans and cisgender men due to the prevalence of Black Pride events and prior evidence indicating that LGBQ Pride events frequently cater to the desires of cisgender men. ^31,41^ To isolate the unique effects of attending racially focused LGBQ Pride events, I controlled for several factors that literature suggests may be associated with LGBQ POC’s psychological wellbeing, including age, income, education, perceived homophobia, religiosity, degree of outness to family, and feelings of connectedness to the LGBQ community/people. ^42-44^

## Methods

### Data and Study Participants

Hypotheses were tested using data from the 2013 Social Justice Sexuality Project (SJSP).^45^. The purpose of SJSP was to examine the sociopolitical experiences unique to LGBQ persons and document the lived experiences of LGBQ people of color ^45^ The SJSP is a cross-sectional national survey administered to Caucasian, Black, Latinx/o/a, Asian, Indigenous, and Multi-Racial American lesbian, gay, bisexual, and adults aged 18 years and older. Participants for the original survey were recruited between January and December 2010 from all 50 US states, Washington, DC, and Puerto Rico through internet and venue-based sampling at LGBQ Pride events, snowball sampling, and respondent-driven sampling methods.

The sample for the present study comprised 3,417 cisgender adults who responded to the SJSP survey (see Table 1 for demographic statistics). The sample was restricted to cisgender adults due to the small sample size of transgender and gender variant individuals as well as prior evidence indicating the presence of additional layers of marginalization at both general LGBQ and racially focused LGBQ that is uniquely experienced by transgender and gender variant individuals. ^46,47^ I further restricted the sample to participants with complete data on all study variables, resulting in an analytic sample size of 2,486. The University of Georgia’s IRB determined the present study to be exempt from review because it is a secondary analysis of existing data.

**Table 1.**
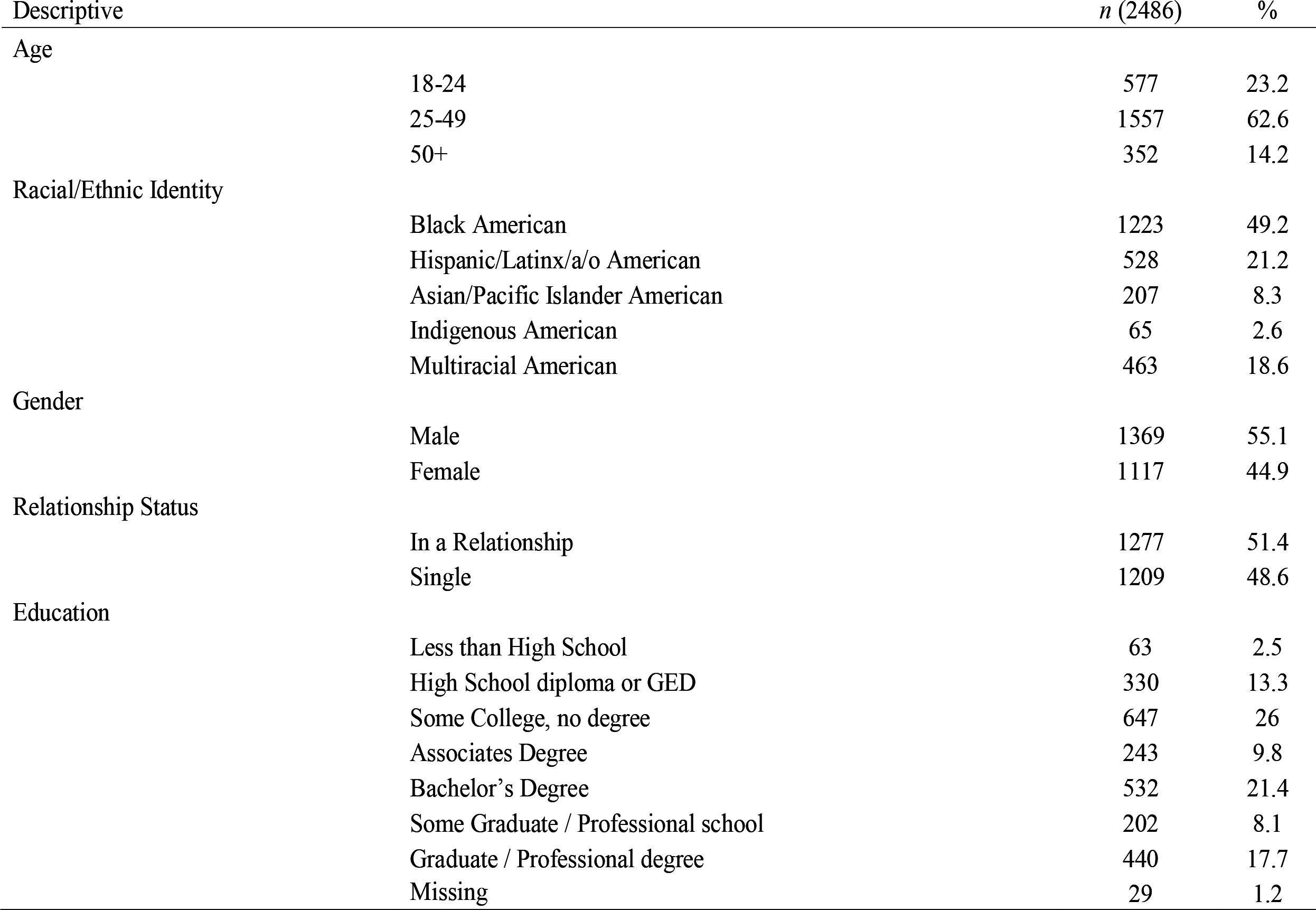
Participant Descriptive Statistics

Participant Descriptive Statistics
| Descriptive |  | *n* (2486) | % |
| --- | --- | --- | --- |
| Age | 18-24 | 577 | 23.2 |
|  | 25-49 | 1557 | 62.6 |
|  | 50+ | 352 | 14.2 |
| Racial/Ethnic Identity | Black American | 1223 | 49.2 |
|  | Hispanic/Latinx/a/o American | 528 | 21.2 |
|  | Asian/Pacific Islander American | 207 | 8.3 |
|  | Indigenous American | 65 | 2.6 |
|  | Multiracial American | 463 | 18.6 |
| Gender | Male | 1369 | 55.1 |
|  | Female | 1117 | 44.9 |
| Relationship Status | In a Relationship | 1277 | 51.4 |
|  | Single | 1209 | 48.6 |
| Education | Less than High School | 63 | 2.5 |
|  | High School diploma or GED | 330 | 13.3 |
|  | Some College, no degree | 647 | 26 |
|  | Associates Degree | 243 | 9.8 |
|  | Bachelor's Degree | 532 | 21.4 |
|  | Some Graduate / Professional school | 202 | 8.1 |
|  | Graduate / Professional degree | 440 | 17.7 |
|  | Missing | 29 | 1.2 |

### Measures

Psychological wellbeing was assessed using a 4-item measure. Each item began with the stem “Over the past week, how often have you felt…” followed by a feeling. Items included (a) that you were just as good as other people, (b) hopeful about the future, (c) happy, and (d) that you enjoyed life. Responses were recorded using a Likert scale ranging from 1 (*never*) to 4 (*most of the time*). Cronbach’s α was .88.

Attending racially focused LGBQ Pride events was assessed using a single indicator, “how often have you attended a racial or ethnic LGBT Pride festival (e.g., Black Pride, Latina/o Pride, Asian Pride, etc.).” Responses were recorded using a Likert scale ranging from 1 (*never*) to 6 (*frequently*).

Participants self-reported their racial identity as Black American, Latinx/o/a American, Asian American, Indigenous American, or Multi-Racial American. Since only cisgender POC were included in this analysis, gender was binary coded as either cisgender male or cisgender female.

#### Covariates

Age was measured in years. Education was measured in years of formal education completed. Total household income in the past year was measured as an ordinal variable in increments of $2,499 (1=<$8500, 2=$8500–10,999, 3=$11,000–13,499, 4=$13,500–14,999, etc.).

Perceived homophobia was assessed using a 3-item measure. Each item began with the stem “Homophobia is a problem within…” followed by an environmental context. Items included (a) racial/ethnic community, (b) neighborhood, and (c) all communities of color. Responses were recorded using a Likert scale ranging from 1 (*strongly disagree*) to 6 (*strongly agree*). Cronbach’s α was .74.

Religiously was assessed using a 5-item assessment of participants’ engagement in spiritual or religious practices. Example items include “I pray daily” and “My faith impacts many of my decisions.” Responses were recorded using a Likert scale ranging from 1 (*strongly disagree*) to 4 (*strongly agree*). Cronbach’s α was .94.

Degree of outness was assessed using a 6-item measure. Each item asked participants to rate how many people with their family, friends, religious community, co-workers, people in their neighborhood, and people online they had disclosed their LGBQ status to. Responses were recorded using a Likert scale ranging from 0 (*none*) to 4 (*all*). Cronbach’s α was .88.

Feelings of connectedness to the LGBQ community/people were assessed using a 3-item measure. Each item began with the stem “I feel…”. Items included (a) connected to my local LGBT community, (b) that the problems faced by the LGBT community are also my problems, and (c) a bond with other LGBT people. Responses were recorded using a Likert scale ranging from 1 (*strongly disagree*) to 6 (*strongly agree*). Cronbach’s α was .77.

### Analytic Strategy

Hypotheses were tested using path analysis in Mplus 8. There were two steps in the analytic procedures in this study. First, a multiple linear regression was conducted to examine the association between attending racially focused LGBQ Pride events and cisgender LGBQ POC’s psychological wellbeing, controlling for all covariates. Second, two multigroup analysis models were applied separately to investigate the modulating effects of racial identity or gender. Each multigroup model was conducted based on the same analytic procedures, which involved comparisons of nested models. First, I established multiple linear regression models wherein all regression coefficients were freely estimated across all groups. Second, I constrained the model’s parameters to be equal across all groups. I conducted a nested chi-square difference test to assess whether holding the coefficients equal significantly worsened the model fit. All covariates were included in each model.

## Results

Descriptive statistics and Pearson correlation coefficients between the study variables are displayed in Table 2. Preliminary results provided initial support to the proposed hypotheses as attending racially focused LGBQ Pride events had a significant and positive correlation with LGBQ POC’s psychological wellbeing.

**Table 2.**
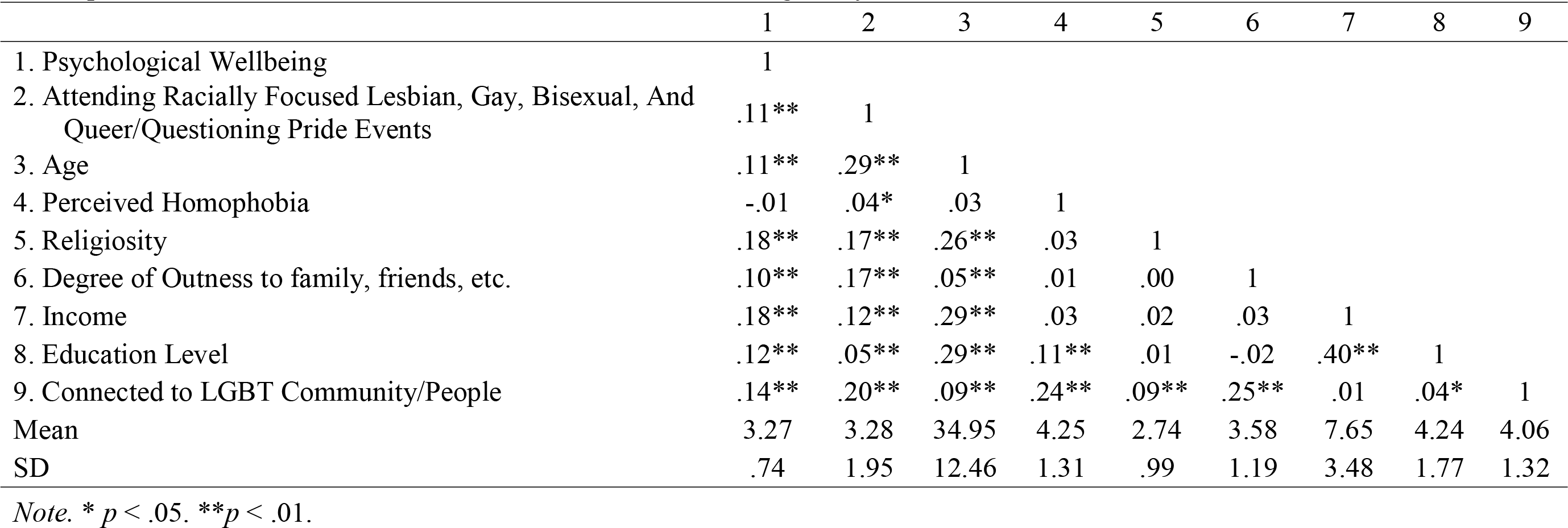
Descriptive Statistics and Two-Tailed Bivariate Correlations Among Study Variables

Table 3 presents the results of the initial multiple linear regression. Consistent with study hypothesis, attending racially focused LGBQ Pride events was positively associated with cisgender LGBQ POC’s psychological wellbeing (β = .04). Additional significant independent variables included: religiosity (β = .16), degree of outness to family (β = .07), income (β = .15), education level (β = .08), and feelings of connectedness to the LGBQ community/people (β = .12).

**Table 3.** Unstandardized Effects of Attending Racially Focused Lesbian, Gay, Bisexual, and Queer/Questioning (LGBQ) Pride Events on LGBQ People Of Color’s Psychological Wellbeing

|  | B | SE | *p* | 95% CI |  |
| --- | --- | --- | --- | --- | --- |
|  |  |  |  | *Lower* | *Upper* |
| Attending Racially Focused Lesbian, Gay, Bisexual, And Queer/Questioning Pride Events | .02* | .01 | .048 | .000 | .031 |
| Age | .00 | .00 | .474 | -.004 | .002 |
| Perceived Homophobia | -.02 | .01 | .059 | -.046 | .000 |
| Religiosity | .12*** | .02 | .000 | .089 | .150 |
| Degree of Outness to family, friends, etc. | .04** | .01 | .002 | .014 | .064 |
| Income | .03*** | .01 | .000 | .022 | .041 |
| Education | .03*** | .01 | .000 | .014 | .049 |
| Feelings of Connectedness to the LGBT Community/People | .07*** | .01 | .000 | .044 | .089 |

*Note.* \* $p < .05$ , \*\* $p < .01$ , \*\*\* $p < .001$ , B = unstandardized beta coefficient, SE = standard error, 95% CI = 95% confidence intervals

Regarding the post hoc multigroup analyses, results from the chi-square difference test that examined potential differences in results based on racial/ethnic groups showed that the hypothesized model was invariant among different racial/ethnic groups, χ^2^_diff(36)_ = 37.57, p = .40, i.e., there was no statistically significant difference in effects between these group. Similar invariant results were found among different gender categories, χ^2^_diff(9)_ = 14.96, p = .09. These results indicate that neither racial/ethnic identity nor gender identity influenced the association between attending racially focused LGBQ Pride events and cisgender LGBQ POC’s psychological wellbeing.

## Discussion

Framed by an intersectional perspective and informed by minority resiliency models, the present study is among the first to investigate the extent to which attending racially focused LGBQ Pride events was associated with cisgender LGBQ POC’s psychological wellbeing. Intersectionality has become increasingly recognized as essential component to investigating mental health disparities experienced by LGBQ adults as studies perpetually demonstrate significant levels of within-group differences in that LGBQ POC often experience higher levels of psychological distress and mental health issues. ^12-15^ Unfortunately, literature investigating culturally relevant promotive factors is largely absent. This study contributes to this nascent body of literature by demonstrating a positive association between attending racially focused LGBQ Pride events was positively and cisgender LGBQ POC’s psychological wellbeing. This positive association that was not modulated by participants’ racial/ethnic identity or gender.

Study findings are consistent with prior theoretical research which indicates that among LGBQ POC attending such events may contribute to their psychological wellbeing by catering to their wants, needs, and desires that go unacknowledged or unaddressed by general LGBQ Pride events. ^48-50^ Results of this study, not only, corroborated prior findings but also indicated that attending racially focused LGBQ Pride events had an equally positive impact on all racial/ethnic groups included in this analysis and cisgender men and women. These findings suggest that large-scale community building events may serve as unique culturally enriching environments wherein cisgender LGBQ POC, regardless of racial/ethnic group or gender, are able to cultivate areas of psychological resilience and wellbeing. These results are notable because the existence of racially focused LGBQ Pride events are frequently critiqued for being *unnecessary* due to the prevalence of general LGBQ Pride events. ^49,51^ Study results indicate that racially focused LGBQ Pride events provide LGBQ POC with unique socioculturally relevant assets that support their psychological wellbeing.

### Limitations

Results of this study should be considered in the context of their limitations. First, this was an analysis of cross-sectional data, which limited my ability to draw causal conclusions; prospective examinations of these associations are needed. Second, data used in this study was from a non-probability sample of LGBQ POC adults and, as such, our findings may not be generalizable to the broader population of LGBQ POC. Third, this study only included cisgender LGBQ POC thus limiting the generalizability of these results to transgender or gender diverse populations. Fifth, all measures included in this analysis were self-report, which are subject to social desirability and recall biases. As such, it will be important to replicate these findings in a longitudinal study with a nationally representative sample, and for future studies to assess other promotive factors that were not captured in this study (e.g., social networks, access to LGBT clinics and centers, and LGBQ POC mentors), given that these factors have been identified as promotive factors in past studies. ^52,53^

## Conclusions

Despite their sociocultural relevance, racially focused LGBQ Pride events are often less funded or supported by non-corporate sponsors, thus limiting their ability to maintain their integrity while expanding their capacity to meet the needs of their intended audience. ^51,54^ Further research is needed to develop a more robust understanding of how attending racially focused LGBQ Pride events may be uniquely beneficial to LGBQ POC’s mental health. Such research would have significant implications for the creation of innovative structural prevention and intervention efforts aimed at addressing the mental health disparities experienced by LGBQ POC.

## Data Availability

All data produced in the present work are contained in the manuscript.

## References

1. Meyer IH, Dietrich J, Schwartz S. Lifetime prevalence of mental disorders and suicide attempts in diverse lesbian, gay, and bisexual populations. Am J Public Health. Jun 2008;98(6):1004–6. doi:10.2105/AJPH.2006.096826

2. O’Donnell S, Meyer IH, Schwartz S. Increased risk of suicide attempts among Black and Latino lesbians, gay men, and bisexuals. American Journal of Public Health. Jun 2011;101(6):1055–1059. doi:10.2105/AJPH.2010.300032

3. Choi K-H, Paul J, Ayala G, Boylan R, Gregorich SE. Experiences of discrimination and their impact on the mental health among African American, Asian and Pacific Islander, and Latino men who have sex with men. American Journal of Public Health. May 2013;103(5):868–874. doi:10.2105/AJPH.2012.301052

4. Chakraborty A, McManus S, Brugha TS, Bebbington P, King M. Mental health of the non-heterosexual population of England. The British journal of psychiatry. 2011;198(2):143–148.

5. Colpitts E, Gahagan J. The utility of resilience as a conceptual framework for understanding and measuring LGBTQ health. International journal for equity in health. 2016;15(1):1–8.

6. Fish JN. Future directions in understanding and addressing mental health among LGBTQ youth. Journal of Clinical Child & Adolescent Psychology. 2020;49(6):943–956.

7. Meyer IH. Prejudice, social stress, and mental health in lesbian, gay, and bisexual populations: conceptual issues and research evidence. Psychol Bull. Sep 2003;129(5):674–697. doi:10.1037/0033-2909.129.5.674

8. Meyer IH. Resilience in the study of minority stress and health of sexual and gender minorities. Psychology of Sexual Orientation and Gender Diversity. 2015;2(3):209.

9. Butler SS. Social networks and social isolation among LGBT older adults. Social isolation of older adults: Strategies to bolster health and well-being. 2019:181–196.

10. Kiperman S, Schacter H, Judge M, DeLong G. A Mixed Methods Investigation of LGBTQ+ Victimization, Its Relationship to LGBTQ+ Identity Development, and the Buffering Potential of Social Support and Outness. 2022;

11. Allen SH, Leslie LA. Considering the role of nativity in the health and psychological wellbeing of Black LGBT adults. Journal of homosexuality. 2018;66(13):1769–1796. doi:10.1080/00918369.2018.1511134

12. Gilbert KL, Ray R, Siddiqi A, et al. Visible and invisible trends in Black men’s health: pitfalls and promises for addressing racial, ethnic, and gender inequities in health. Annual Review of Public Health. 2016;37:295–311. doi:10.1146/annurev-publhealth-032315-021556

13. Seaton EK, Caldwell CH, Sellers RM, Jackson JS. An intersectional approach for understanding perceived discrimination and psychological well-being among African American and Caribbean Black youth. Developmental psychology. 2010;46(5):1372.

14. Tsang EY-h, Qiao S, Wilkinson JS, Fung AL-c, Lipeleke F, Li X. Multilayered stigma and vulnerabilities for HIV infection and transmission: A qualitative study on male sex workers in zimbabwe. American Journal of Men’s Health. Jan-Feb 2019;13(1):1557988318823883. doi:10.1177/1557988318823883

15. Vu M, Li J, Haardorfer R, Windle M, Berg CJ. Mental health and substance use among women and men at the intersections of identities and experiences of discrimination: insights from the intersectionality framework. article. BMC Public Health. Jan 23 2019;19(1):108. doi:10.1186/s12889-019-6430-0

16. Huang Y-P, Brewster ME, Moradi B, Goodman MB, Wiseman MC, Martin A. Content analysis of literature about LGB people of color: 1998-2007. The Counseling Psychologist. 2010;38(3):363–396.

17. Toomey RB, Huynh VW, Jones SK, Lee S, Revels-Macalinao M. Sexual minority youth of color: A content analysis and critical review of the literature. Journal of gay & lesbian mental health. 2017;21(1):3–31. doi:10.1080/19359705.2016.1217499

18. Ryan C, Huebner D, Diaz RM, Sanchez J. Family rejection as a predictor of negative health outcomes in white and Latino lesbian, gay, and bisexual young adults. Pediatrics. 2009;123(1):346–352.

19. Moradi B, DeBlaere C, Huang Y-P. Centralizing the experiences of LGB people of color in counseling psychology 1Ψ7. The Counseling Psychologist. 2010;38(3):322–330.

20. Sadika B, Wiebe E, Morrison MA, Morrison TG. Intersectional microaggressions and social support for LGBTQ persons of color: A systematic review of the Canadian-based empirical literature. Journal of GLBT family studies. 2020;16(2):111–147.

21. Loeb AJ, Wardell D, Johnson CM. Coping and healthcare utilization in LGBTQ older adults: A systematic review. Geriatric Nursing. Jul-Aug 2021;42(4):833–842. doi:10.1016/j.gerinurse.2021.04.016

22. Chang CJ, Feinstein BA, Meanley S, Flores DD, Watson RJ. The Role of LGBTQ Identity Pride in the Associations among Discrimination, Social Support, and Depression in a Sample of LGBTQ Adolescents. Annals of LGBTQ Public and Population Health. 2021;2(3):203–219.

23. Bridge L, Smith P, Rimes KA. Self-esteem in sexual minority young adults: a qualitative interview study exploring protective factors and helpful coping responses. International Review of Psychiatry. 2022:1–9.

24. Stokols D. Translating social ecological theory into guidelines for community health promotion. American journal of health promotion. 1996;10(4):282–298.

25. Cote M, Nightingale AJ. Resilience thinking meets social theory: situating social change in socio-ecological systems (SES) research. Progress in Human Geography. 2012;36(4):475–489.

26. Irwin JA, Coleman JD, Fisher CM, Marasco VM. Correlates of suicide ideation among LGBT Nebraskans. Journal of homosexuality. 2014;61(8):1172–1191. doi:10.1080/00918369.2014.872521

27. Conover KJ, Israel T. Microaggressions and social support among sexual minorities with physical disabilities. Rehabilitation psychology. May 2019;64(2):167. doi:10.1037/rep0000250

28. Brownfield JM, Brown C, Jeevanba SB, VanMattson SB. More than simply getting bi: An examination of coming out growth for bisexual individuals. Psychology of Sexual Orientation and Gender Diversity. 2018;5(2):220. doi:10.1037/sgd0000282

29. Armstrong EA, Crage SM. Movements and memory: The making of the Stonewall myth. American sociological review. 2006;71(5):724–751. doi:10.1177/000312240607100502

30. Browne K. A party with politics?(Re) making LGBTQ Pride spaces in Dublin and Brighton. Social & Cultural Geography. 2007;8(1):63–87. doi:10.1080/14649360701251817

31. Kenttamaa Squires K. Rethinking the homonormative? Lesbian and Hispanic Pride events and the uneven geographies of commoditized identities. Social & Cultural Geography. 2019;20(3):367–386. doi:10.1080/14649365.2017.1362584

32. Becker AB, Copeland L. Networked publics: How connective social media use facilitates political consumerism among LGBT Americans. Journal of Information Technology & Politics. 2016;13(1):22–36. doi:10.1080/19331681.2015.1131655

33. Ciszek EL. Identity, culture, and articulation: A critical-cultural analysis of strategic LGBT advocacy outreach. University of Oregon; 2014. https://www.proquest.com/dissertations-theses/identity-culture-articulation-critical-cultural/docview/1619367520/se-2

34. Olson ED. An exploration of lesbian, gay, bisexual, and transgender pride festival sponsors. Taylor & Francis; 2017:60–73.

35. Csensich CM. Consumerism and Pride: The Fate of Paris’ Marais “Gayborhood”. University of South Florida; 2021. https://www.proquest.com/dissertations-theses/consumerism-pride-fate-paris-marais-gayborhood/docview/2524415086/se-2?accountid=14537

36. Puar J. A transnational feminist critique of queer tourism. Antipode. 2002;34(5):935–946. doi:10.1111/1467-8330.00283

37. Hunter MA. All the gays are White and all the Blacks are straight: Black gay men, identity, and community. Sexuality Research and Social Policy. 2010;7(2):81–92. doi:10.1007/s13178-010-0011-4

38. Walsh CF. “It really is not just gay, but African American gay”: the impact of community and church on the experiences of black lesbians living in North Central Florida. Journal of Homosexuality. Sep 2016;63(9):1236–1252. doi:10.1080/00918369.2016.1151694

39. Brathwaite LF. Why Black Pride Matters. Advocate. November 25, 2019. https://www.advocate.com/current-issue/2016/4/28/why-black-pride-matters

40. Bartone MD. “Nothing has stopped me. I keep going:” Black gay narratives. Journal of LGBT Youth. 2017;14(3):317–329. doi:10.1080/19361653.2017.1324342

41. Jeppesen S. Queer anarchist autonomous zones and publics: Direct action vomiting against homonormative consumerism. SEXUALITIES. 2010;13(4):463–478. doi:10.1177/1363460710370652

42. Dürrbaum T, Sattler FA. Minority stress and mental health in lesbian, gay male, and bisexual youths: A meta-analysis. Journal of LGBT Youth. 2020;17(3):298–314. doi:10.1080/19361653.2019.1586615

43. Carvalho SA, Guiomar R. Self-Compassion and Mental Health in Sexual and Gender Minority People: A Systematic Review and Meta-Analysis. LGBT health. Mar 29 2022;doi:10.1089/lgbt.2021.0434

44. Kaniuka A, Pugh KC, Jordan M, et al. Stigma and suicide risk among the LGBTQ population: Are anxiety and depression to blame and can connectedness to the LGBTQ community help? Journal of Gay & Lesbian Mental Health. 2019;23(2):205–220. doi:10.1080/19359705.2018.1560385

45. Battle J, Pastrana AJ, Daniels J. Data from: Social Justice Sexuality Project: 2010 National Survey, including Puerto Rico. 2013. doi:10.3886/ICPSR34363.v1

46. McCartan A, Nash CJ. Creating queer safe space: relational space-making at a grassroots LGBT pride event in Scotland. Gender, Place & Culture. 2022:1–21. doi:10.1080/0966369X.2022.2052019

47. Enguix B. Identities, sexualities and commemorations: Pride parades, public space and sexual dissidence. Anthropological Notebooks. 2009;15(2)

48. Squires KK. Declining Gayborhood or Homonormative Playground in the Making? South Beach Reinvented. University of Miami; 2014.

49. Baez R. Why We Need’Reclaim Pride’. The Gay & Lesbian Review Worldwide. 2019;26(3):22–26.

50. McNeill Z, Smith K. Whose Pride Is This Anyway? The Quare Performance of the# Black Pride4. The Palgrave Handbook of Queer and Trans Feminisms in Contemporary Performance. Springer; 2021:203–222.

51. Smith CG. “Where u from, who u wit?!” Black Pride Festivals as Itinerant Hospitality. Journal of Canadian Studies. 2020;54(2-3):395–414. doi:10.3138/jcs-2019-0039

52. Kim H-J, Fredriksen-Goldsen KI, Bryan AE, Muraco A. Social network types and mental health among LGBT older adults. The Gerontologist. 2017;57(suppl_1):S84–S94.

53. Hellman RE, Klein E. A program for lesbian, gay, bisexual, and transgender individuals with major mental illness. Journal of Gay & Lesbian Psychotherapy. 2004;8(3-4):67–82.

54. Duggan L. The twilight of equality?: Neoliberalism, cultural politics, and the attack on democracy. Beacon Press; 2012.

